# BNT162b2 induces SARS-CoV-2-neutralising antibodies and T cells in humans

**DOI:** 10.1101/2020.12.09.20245175

**Authors:** Ugur Sahin, Alexander Muik, Isabel Vogler, Evelyna Derhovanessian, Lena M. Kranz, Mathias Vormehr, Jasmin Quandt, Nicole Bidmon, Alexander Ulges, Alina Baum, Kristen Pascal, Daniel Maurus, Sebastian Brachtendorf, Verena Lörks, Julian Sikorski, Peter Koch, Rolf Hilker, Dirk Becker, Ann-Kathrin Eller, Jan Grützner, Manuel Tonigold, Carsten Boesler, Corinna Rosenbaum, Ludwig Heesen, Marie-Cristine Kühnle, Asaf Poran, Jesse Z. Dong, Ulrich Luxemburger, Alexandra Kemmer-Brück, David Langer, Martin Bexon, Stefanie Bolte, Tania Palanche, Armin Schultz, Sybille Baumann, Azita J. Mahiny, Gábor Boros, Jonas Reinholz, Gábor T. Szabó, Katalin Karikó, Pei-Yong Shi, Camila Fontes-Garfias, John L. Perez, Mark Cutler, David Cooper, Christos A. Kyratsous, Philip R. Dormitzer, Kathrin U. Jansen, Özlem Türeci

## Abstract

BNT162b2, a lipid nanoparticle (LNP) formulated nucleoside-modified messenger RNA (mRNA) encoding the severe acute respiratory syndrome coronavirus 2 (SARS-CoV-2) spike protein (S) stabilized in the prefusion conformation, has demonstrated 95% efficacy to prevent coronavirus disease 2019 (COVID-19). Recently, we reported preliminary BNT162b2 safety and antibody response data from an ongoing placebo-controlled, observer-blinded phase 1/2 vaccine trial^1^. We present here antibody and T cell responses from a second, non-randomized open-label phase 1/2 trial in healthy adults, 19-55 years of age, after BNT162b2 prime/boost vaccination at 1 to 30 µg dose levels. BNT162b2 elicited strong antibody responses, with S-binding IgG concentrations above those in a COVID-19 human convalescent sample (HCS) panel. Day 29 (7 days post-boost) SARS-CoV-2 serum 50% neutralising geometric mean titers were 0.3-fold (1 µg) to 3.3-fold (30 µg) those of the HCS panel. The BNT162b2-elicited sera neutralised pseudoviruses with diverse SARS-CoV-2 S variants. Concurrently, in most participants, S-specific CD8^+^ and T helper type 1 (T_H_1) CD4^+^ T cells had expanded, with a high fraction producing interferon-γ (IFNγ). Using peptide MHC multimers, the epitopes recognised by several BNT162b2-induced CD8^+^ T cells when presented on frequent MHC alleles were identified. CD8^+^ T cells were shown to be of the early-differentiated effector-memory phenotype, with single specificities reaching 0.01-3% of circulating CD8^+^ T cells. In summary, vaccination with BNT162b2 at well tolerated doses elicits a combined adaptive humoral and cellular immune response, which together may contribute to protection against COVID-19.

## Introduction

Given the high impact of the pandemic caused by the severe acute respiratory syndrome coronavirus 2 (SARS-CoV-2) on human society, the rapid development of safe and effectively prophylactic vaccines is of utmost importance.

Lipid nanoparticle (LNP) formulated messenger RNA (mRNA) vaccine technology delivers the precise genetic information of the immunogen to antigen presenting cells and elicits potent immune responses^2^. mRNA is transiently expressed, does not integrate into the genome, and is degraded by physiological pathways. mRNA vaccines are molecularly well defined, and are synthesized efficiently from DNA templates by *in vitro* transcription, which is cell- and animal-origin material-free^3–5^. mRNA production and LNP formulation are fast processes of high scalability, rendering this technology suitable for rapid vaccine development and pandemic vaccine supply^6–8^.

Within ‘Project Lightspeed’, the joint BioNTech-Pfizer COVID-19 RNA vaccine development program, two phase 1/2 umbrella trials, one in Germany (NCT04380701) and one in the USA (NCT04368728) investigate a total of four RNA-LNP vaccine candidates in. Recently, we reported preliminary clinical data from these studies on the two most advanced candidates BNT162b1^9,10^ and BNT162b2^1^. Each candidate is an LNP-formulated, pharmacologically optimized^11,12^, N^1^-methylpseudouridine (m1Ψ) nucleoside-modified mRNA (modRNA)^13^ administered intramuscularly as a prime-boost 21 days apart. BNT162b1 encodes a trimerized, secreted version of the receptor-binding domain (RBD) of S, whereas BNT162b2 encodes the full-length SARS-CoV-2 S stabilised in the prefusion conformation (P2 S)^14^.

BNT162b2 in a 30 µg two dose regimen has been selected for advancement into an ongoing phase 2/3 trial, guided by the totality of data obtained in the two phase 1/2 trials and NHP challenge studies^1,15^. In the placebo-controlled, observer-blind USA phase 1/2 trial, immunisation of 18-55 and 65-83 year old participants with BNT162b2 at dose levels of up to 30 µg was associated with generally mild to moderate local injection site reactions and systemic events such as fatigue, headache, and myalgia^1^. BNT162b2 elicited robust S1-binding immunoglobulin G (IgG) concentrations and SARS-CoV-2 neutralising titers. Geometric mean 50% neutralising titers (GMTs) of sera drawn from younger and older adults seven days after the second dose of 30 µg BNT162b2 were 3.8-fold and 1.6-fold, respectively, the GMT of a panel of COVID-19 convalescent human sera. Complementing and expanding the published findings in the USA phase 1/2 trial, we now provide data.from the German trial (NCT04380701, EudraCT: 2020-001038-36). We report immunogenicity and safety of prime-boost vaccination with 1, 10, 20 and 30 µg BNT162b2 in participants 19-55 years of age, including neutralising antibody GMTs up to day 85 after the first dose (approximately two months after the booster dose) and detailed characterisation of T cell responses, including the first identification of epitopes recognised by CD8^+^ T cells induced by a COVID-19 vaccine.

### Study design and analysis sets

Participants 19-55 years of age were vaccinated with BNT162b2 in Germany (Extended Data Fig. 1). Participants’ mean age was 40 years; 56% were female, and all were Caucasian (Extended Data Table 1). Twelve participants per dose cohort were assigned to receive a priming dose of 1, 10, 20 or 30 μg on day 1 and a booster dose on day 22 (Extended Data Table 2). One individual each in the 1 µg dose cohort and the 10 µg dose cohort discontinued prior to the boost. In each dose group, antibody levels and virus neutralisation titers were assessed in 10 to 12 participants per timepoint (up to day 85, 63 days post-boost), and peripheral blood mononuclear cells (PBMCs) from 8 to 10 participants were analysed for cellular immune responses at baseline and day 29 (7 days post-boost) (Extended Data Table 2).

### Safety and tolerability

Briefly, no serious adverse events (SAE) and no withdrawals due to related adverse events (AEs) were observed at any dose level. Local reactions, predominantly pain at the injection site, were mild to moderate (grade 1 and 2) and were similar in frequency and severity after the priming and booster doses (Extended Data Fig. 2a and Extended Data Table 3a). The most common systemic AEs were fatigue followed by headache and only two participants reported fever, which was mild (Extended Data Fig. 2b and Extended Data Table 3b). Transient chills were more common after the boost, dose-dependent, and occasionally severe. Muscle pain and joint pain were also more common after the boost and showed dose-dependent severity. There were no grade 4 reactions. Generally, reactions had their onset within 24 hours of immunisation, peaked on the day after immunisation, and mostly resolved within 2-3 days. Reactions did not require treatment or could be managed with simple measures (*e*.*g*. paracetamol).

No clinically significant changes in routine clinical laboratory values occurred after BNT162b2 vaccination. In line with previous reports for RNA-based vaccines^1,9,10,16^, a mild drop of blood lymphocyte counts (without concomitant neutropenia) and an increase in C-reactive protein (CRP) were observed, both transient, dose-dependent and within or close to laboratory normal levels (Extended Data Fig. 3). Both effects are considered pharmacodynamic markers for the mode-of-action of RNA vaccines: blood lymphocyte counts transiently decrease as the lymphocytes redistribute into lymphoid tissues in response to innate immune stimulation^17^, and CRP is a downstream effect of innate immune modulation^18–21^.

### Characterization of vaccine-induced antibody response

S1- and RBD-binding IgG concentrations and SARS-CoV-2 neutralising titers were assessed at baseline (day 1), 7 and 21 days after the BNT162b2 priming dose (days 8 and 22), and 7, 21, 28 and 63 days after the booster dose (days 29, 43 and 50; day 85 for all dose levels except 1 µg) (Fig. 1, Extended Data Fig. 4, Extended Data Table 2).

**Figure 1.**
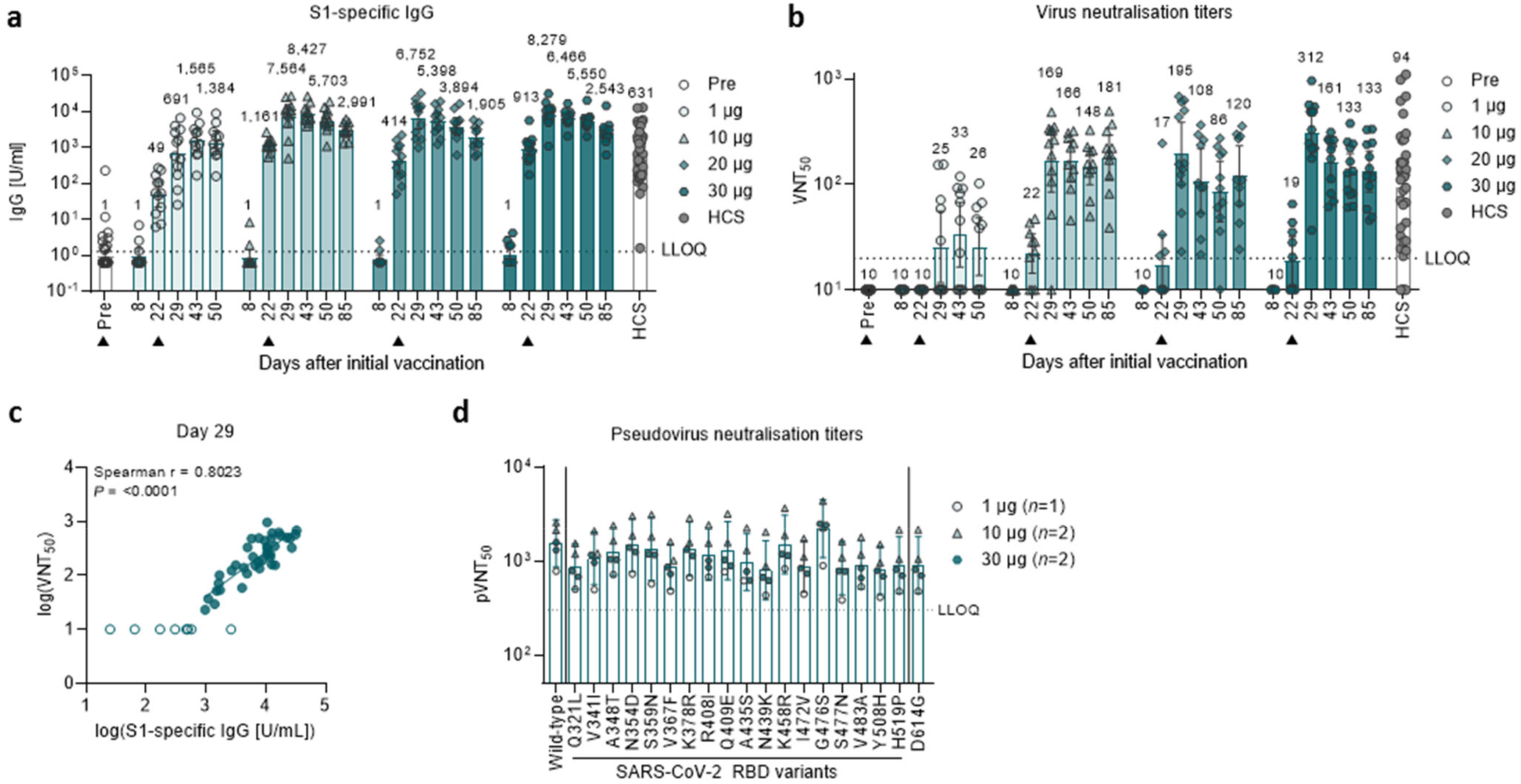
BNT162b2-induced IgG concentrations and virus neutralisation titers. Vaccination schedule and serum sampling are described in Extended Data Fig. 1. Participants were immunised with BNT162b2 on days 1 and 22 (*n*=12 per dose cohort; from day 22 onwards *n*=11 for the 1 µg and 10 µg dose cohorts). Arrowheads indicate days of vaccination. Pre-dose responses across all dose levels were combined. COVID-19 human convalescent samples (HCS, *n*=38) were obtained at least 14 days after PCR-confirmed diagnosis and at a time when the donors were no longer symptomatic. Each serum was tested in duplicate and geometric mean concentrations (GMCs) (**a**) and titers (GMTs) (**b, e**) were plotted. For values below the lower limit of quantification (LLOQ; 1.27 [**a**], 20 [**b**], 300 [**c**]), LLOQ/2 values were plotted. Group GMCs or GMTs (values above bars) with 95% confidence interval. **a**, Recombinant S1-binding IgG GMC. **b**, SARS-CoV-2 50% neutralisation titers (VNT_50_) in immunised participants and HCS. **c**, Nonparametric Spearman correlation of recombinant S1-binding IgG GMCs (as in [**a**]) with VNT_50_ from day 29 sera (as in [**b**]) with data points for participants with GMCs and GMTs below the LLOQ (open circles) excluded. **d**, Pseudovirus 50% neutralisation titers (pVNT_50_) across a pseudovirus panel displaying 19 SARS-CoV-2 S variants including 18 RBD mutants and the dominant S variant D614G (dose levels 10, 30 and 50 µg, *n*=1-2 representative sera each; day 29).

The vaccine elicited strong antibody responses. Twenty-one days after the priming dose, geometric mean concentrations (GMCs) of S1-binding IgG had increased in all dose cohorts, with S1-binding IgG GMCs in the range of 49-1,161 U/mL and evidence of a dose level-dependent response only between the 1 μg and 10 μg dose levels (Fig. 1a). Seven days after the booster dose (day 29), S1-binding IgG GMCs showed a strong booster response ranging from 691-8,279 U/mL. Antibody levels decreased over time, but with S1-binding antibody GMCs still in the range of 1,384-2,991 U/mL at day 85 (63 days after the boost), and hence well above that observed in a panel of sera from SARS-CoV-2 convalescent patients (631 U/mL). Similar observations were made using only the RBD domain as the target antigen (Extended Data Fig. 4).

SARS-CoV-2 50% neutralising geometric mean titers (GMTs) increased modestly and only in a proportion of participants after the priming dose of BNT162b2 (Fig. 1b). By seven days after the booster dose, neutralising GMTs had increased substantially to 169, 195 or 312 in participants immunised with 10 µg, 20 µg or 30 µg BNT162b2, respectively. The 1 µg dose level elicited only a minimal neutralizing response (GMT of 25 at seven days after the boost). On day 43 (21 days after the boost), participants vaccinated with BNT162b2 dose levels between 10 and 30 µg had virus neutralizing GMTs between 108 and 166. Importantly, SARS-CoV-2 neutralising GMTs remained stable up to day 85 (63 days after the boost) with titers ranging from 120 to 181, and thus were 1.3-to 1.9-fold the convalescent serum panel neutralising GMT of 94.

S1-binding IgG GMCs after the boost showed a gradual decline, which is typical of the pattern of proliferation followed by contraction of B cells cognately activated by either natural infection or vaccination^25^. In contrast, GMTs initially decreased after the boost but stabilized around day 43, which implies selection and affinity maturation of functional antibodies.

Neutralising antibody GMTs correlated strongly with S1-binding IgG GMCs (Fig. 1c). In summary, neutralizing responses and antigen-binding IgG responses elicited by BNT162b2 in this study largely mirrored those observed in the U.S.A. study, and for the first time cover extended follow-up until day 85.

A panel of 18 SARS-CoV-2 RBD variants identified through publicly available information^24^ and the dominant non-RBD S variant D614G^25^ were evaluated as targets in pseudovirus neutralisation assays. Sera collected seven days after the booster dose of BNT162b2 showed high neutralising titers to each of the SARS-CoV-2 S variants (Fig. 1d), demonstrating the breadth of the neutralising response against circulating strains.

### Prevalence and magnitude of vaccine-induced T cell responses

T cell responses of 37 BNT162b2 immunised participants from whom sufficient peripheral blood mononuclear cells (PBMCs) were available were analysed pre-vaccination (day 1) and seven days after the booster dose (day 29) by direct ex vivo IFNγ enzyme-linked immunosorbent spot (ELISpot) assay (Fig. 2, Extended Data Fig. 5, Extended Data Table 2). SARS-CoV-2 S is composed of a signal peptide (aa 1-13), the N-terminal S1 protease fragment (aa 14-685), and the C-terminal S2 protease fragment (aa 686-1273). S1 contains the RBD (aa 319-541), which binds to the host receptor, and S2 mediates fusion between the viral envelope and cell membrane. To deconvolute reactivity against S, CD4^+^ or CD8^+^ T cell effectors were stimulated overnight with overlapping peptides representing different portions of the wild-type sequence of SARS-CoV-2 S, namely N-terminal pools ‘S pool 1’ (aa 1-643) and ‘RBD’ (aa 1-16 fused to aa 327-528 of S), and the C-terminal ‘S pool 2’ (aa 633-1273).

**Figure 2.**
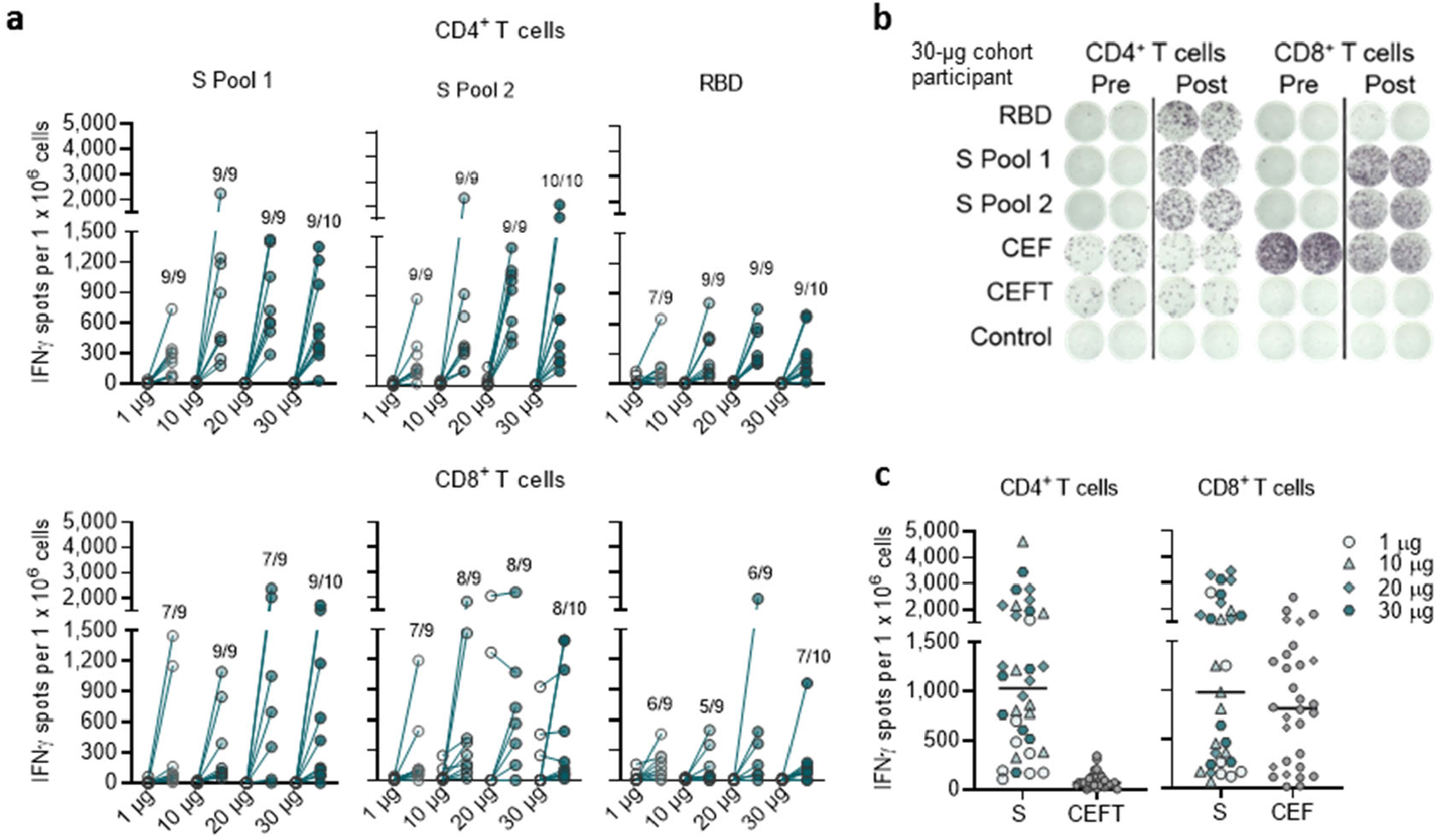
Incidence and magnitude of BNT162b2-induced T cell responses. PBMCs obtained on day 1 (pre-prime) and day 29 (7 days post-boost) (dose cohorts 1, 10 and 20 µg, *n*=9 each; 30 µg, *n*=10) were enriched for CD4^+^ or CD8^+^ T cell effectors and separately stimulated over night with three overlapping peptide pools representing different portions of the wild-type sequence of SARS-CoV-2 S (N-terminal pools S pool 1 and RBD, and the C-terminal S pool 2), for assessment in direct ex vivo IFNγ ELISpot. Common pathogen T cell epitope pools CEF (immune dominant HLA class I epitopes of CMV, EBV, influenza virus) and CEFT (immune dominant HLA class II epitopes CMV, EBV, influenza virus, tetanus toxoid) were used as controls. Cell culture medium served as negative control. Each dot represents the normalised mean spot count from duplicate wells for one study participant, after subtraction of the medium-only control (**a, c**). **a**, Antigen-specific CD4^+^ and CD8^+^ T cell responses for each dose cohort. The number of participants with a detectable T cell response on day 29 over the total number of tested participants per dose cohort is provided. Spot count data from two participants from the 20 µg dose cohort could not be normalised and are not plotted. **b**, Example of CD4^+^ and CD8^+^ ELISpot for a 30 µg dose cohort participant. **c**, S-specific T cell responses in all participants who recognised either S peptide pool and their baseline CEFT- and CEF-specific T cell responses. Horizontal bars indicate median values.

Seven days after the boost with BNT162b2 at any of the doses, robustly expanded SARS-CoV-2 S-specific CD4^+^ T cells were detectable in all 37 participants (Fig. 2a, Extended Data Fig. 5a). In 34 of these participants, comparison to pre-vaccination PBMCs was possible. Thirty of the 34 subjects (88.2%) had de novo (not existent at baseline) CD4^+^ T cell responses against both S pool 1 and S pool 2 of SARS-CoV-2. One participant had de novo response only against S pool 2. The remaining three participants had de novo responses against S pool 1 and low numbers of pre-existing S pool 2-reactive CD4^+^ T cells. In two of these three participants, the pre-existing responses against S pool 2 were amplified by vaccination (from 91 and 188 spots/10^6^ cells pre-vaccination to 1391 and 965 spots after vaccination, respectively), whereas in one of the three participants, the pre-existing responses against S pool 2 remained stable (53 to 140 spots/10^6^ cells). In conclusion, these data demonstrate that in 94.1% (32/34) of participants, two doses of BNT162b2 induce poly-epitopic CD4^+^ T cell responses (de novo or amplified) directed against both N- and C-terminal portions of S and thus against epitopes outside the RBD (Extended Data Fig. 5b).

Although for dose levels ≥10 µg the magnitude of CD4^+^ T cell responses did not appear to be dose-dependent, it varied between individuals. In the strongest responders, the S-specific CD4^+^ T cell responses were more than 10-fold of the individual memory responses to common viruses and recall antigens (those from cytomegalovirus, Epstein Barr virus, influenza virus and tetanus toxoid) (Fig. 2b, c).

Vaccine-induced S-specific CD8^+^ T cell responses were detected in 34 of 37 vaccinated participants (91.9%). The majority were strong responses (Fig. 2a, Extended Data Fig. 5a) comparable to individual memory responses against cytomegalovirus (CMV), Epstein Barr virus (EBV)and influenza virus (Fig. 2b, c). De novo S -specific CD8^+^ T cell responses were induced in 33 participants, these were either directed against both (22 participants), or one of the S pools (S pool 1 in ten participants, and S pool 2 in two participants), indicating a preponderance of a poly-epitopic response including non-RBD S-specific T cells (Extended Data Fig. 5b). In seven participants, pre-existing CD8^+^ T cell responses to S pool 2 were detected that were not further augmented by vaccination. Six out of these seven participants had a concurrent de novo response to pool 1 of S, which in strength did not differ significantly from those observed in individuals without pre-existing responses to S pool 2 (Extended Data Fig. 5c). Of note, the strongest responses (higher than third quartile) against S pool 1 among the 34 participants with detectable CD8^+^ T cell responses were observed in those without pre-existing S pool 2-specific responses.

The magnitude of S-specific CD4^+^ T cell responses correlated positively with S1-binding IgG (Extended Data Fig. 6a), and, in line with the concept of intramolecular help^26^, also with the strength of S-specific CD8^+^ T cell responses (Extended Data Fig. 6b). S-specific CD8^+^ T cell responses also correlated positively with S1-binding IgG (Extended Data Fig. 6c), indicating a convergent development of the humoral and cellular adaptive immunity.

### Polarisation of vaccine-induced T cell responses

To assess functionality and polarisation of S-specific T cells, cytokines secreted in response to stimulation with S pool 1, S pool 2 and RBD pool were determined by intracellular staining (ICS) for IFNγ, IL-2 and IL-4 specific responses in pre- and post-vaccination PBMCs of 37 BNT162b2-immunised participants (Extended Data Table 2). A considerable fraction of vaccine-induced, S-specific CD4^+^ T cells secreted IFNγ, IL-2, or both, while T cells secreting the T_H_2 cytokine IL-4 were barely detectable (Fig. 3a-c, Extended Data Fig. 5d, e). Vaccine-induced S-specific CD8^+^ T cells secreted predominantly IFNγ and lower levels of IL-2 in response to S pool 1 and S pool 2 stimulation. Fractions of IFNγ^+^ CD8^+^ T cells specific to S pool 1 constituted up to about 1% of total peripheral blood CD8^+^ T cells (Fig. 3d). Of note, several of the analysed participants (*n*=3 in the 20 µg dose cohort and *n*=3 in the 30 µg dose cohort) displayed pre-existing S pool 2 specific CD8^+^ T cell responses, which in 5 out of the 6 participants were not further amplified after vaccination. A strong pre-existing S pool 2 specific IFNγ^+^ CD4^+^ T cell response was detectable in one participant (20 µg dose cohort) (Fig. 3c).

**Figure 3.**
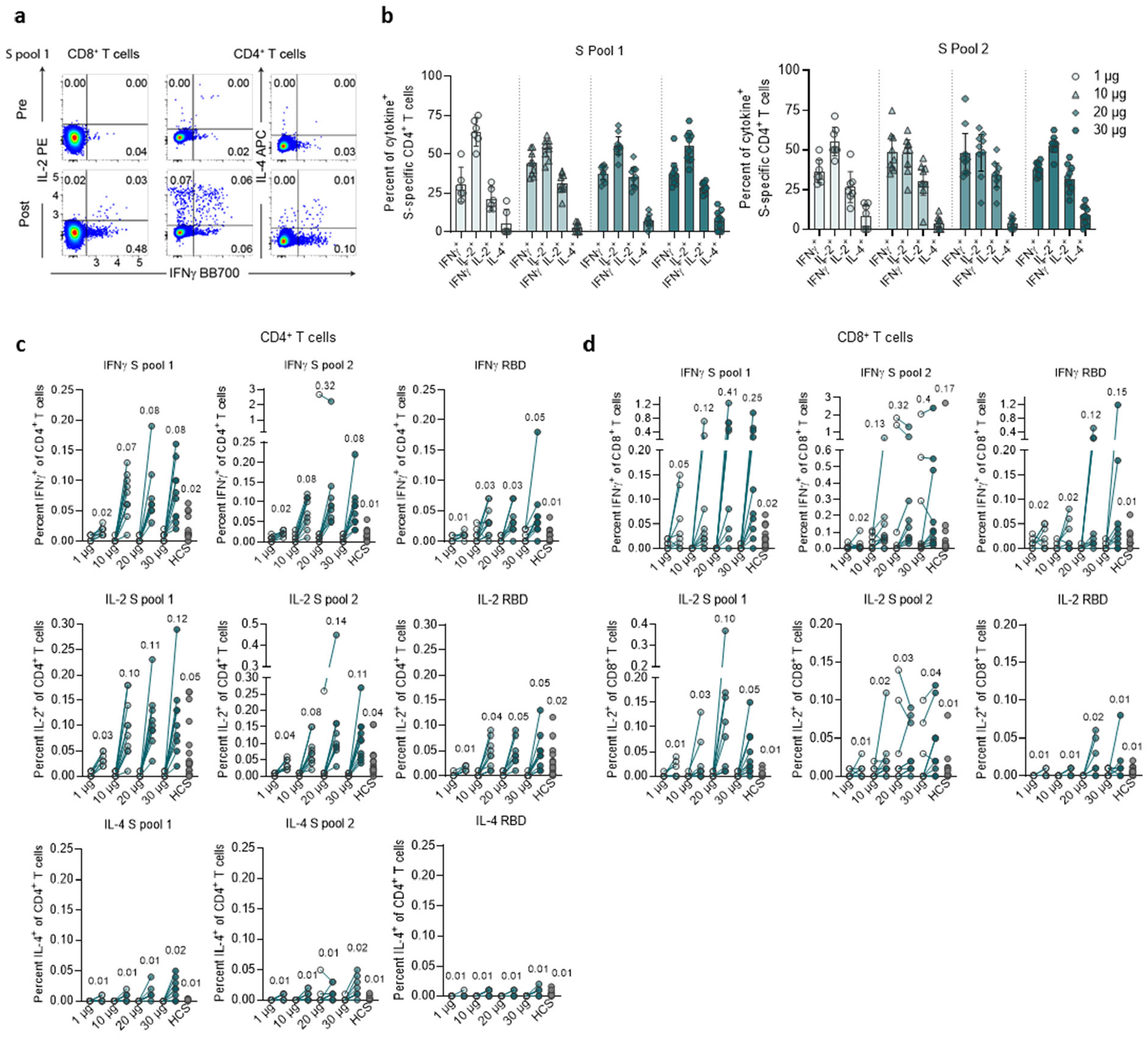
Cytokine polarisation of BNT162b2-induced T cells. PBMCs obtained on day 1 (pre-prime) and day 29 (7 days post-boost) (dose cohorts 1 µg, *n*=8; 10 and 30 µg, *n*=10 each; 20 µg, *n*=9) and COVID-19 recovered donors (HCS, *n*=18; **c, d**) were stimulated over night with three overlapping peptide pools representing different portions of the wild-type sequence of SARS-CoV-2 S (N-terminal pools S pool 1 [aa 1-643] and RBD [aa1-16 fused to aa 327-528 of S], and the C-terminal S pool 2 [aa 633-1273]), and analysed by flow cytometry (for gating strategy see Supplementary Fig. 1). **a**, Example of pseudocolor flow cytometry plots of cytokine-producing CD4^+^ and CD8^+^ T cells from a 30 µg dose cohort participant in response to S pool 1. **b**, S-specific CD4^+^ T cells producing the indicated cytokine as a fraction of total cytokine-producing S-specific CD4^+^ T cells in response to S pool 1 and S pool 2. CD4 non-responders (<0.03% total cytokine producing T cells: 1 µg, *n*=2 [S pool 1] and *n*=1 [S pool 2]; 10 µg, *n*=1) were excluded. Arithmetic mean with 95% confidence interval. **c**, S-specific CD4^+^ (S pool 1, S pool 2 and RBD) and **d**, CD8^+^ T cells (S pool 1, S pool 2 and RBD) producing the indicated cytokine as a fraction of total circulating T cells of the same subset. Values above data points indicate mean fractions per dose cohort. Participant PBMCs were tested as single instance (**b-d**).

In both assay systems, cytokine production of CD4^+^ as well as CD8^+^ T cells in response to peptide pools comprising the full SARS-CoV-2 S exceeded the responses against the RBD peptide pool, further confirming the poly-epitopic nature of T cell responses elicited by BNT162b2. The mean fraction of BNT162b2-induced S-specific IFNγ^+^ or IL-2^+^ CD4^+^ and CD8^+^ T cells within total circulating T cells was higher than that detected in eighteen control subjects who had recovered from COVID-19 (HCS) (Fig. 3c, d).

### Epitope specificity and phenotype of CD8^+^ T cells

CD8^+^ T cell responses were characterised on the epitope level in three BNT162b2 vaccinated participants. To this aim, pre- and post-vaccination PBMCs were stained with individualised peptide/MHC multimer staining cocktails for flow cytometry analysis. Twenty-three (4 for HLA-B*0702, 19 for HLA-A*2402), 14 (HLA-B*3501) and 23 (7 for HLA-B*4401, 16 for HLA-A*0201) diverse peptide/MHC allele pairs were used for participants 1, 2 and 3, respectively. This approach identified de novo induced CD8^+^ T cell reactivities against multiple epitopes for each participant, adding up to a total of eight different epitope/MHC pairs spread across the full length of S (Fig. 4a, c). The magnitude of epitope-specific T cell responses ranged between 0.01-3.09% of peripheral CD8^+^ T cells, and the most profound expansion was observed for HLA-A*0201 YLQPRTFLL (3.09% multimer^+^ of CD8^+^), HLA-A*2402 QYIKWPWYI (1.27% multimer^+^ of CD8^+^) and HLA-B*3501 QPTESIVRF (0.17% multimer^+^ of CD8^+^).

**Figure 4.**
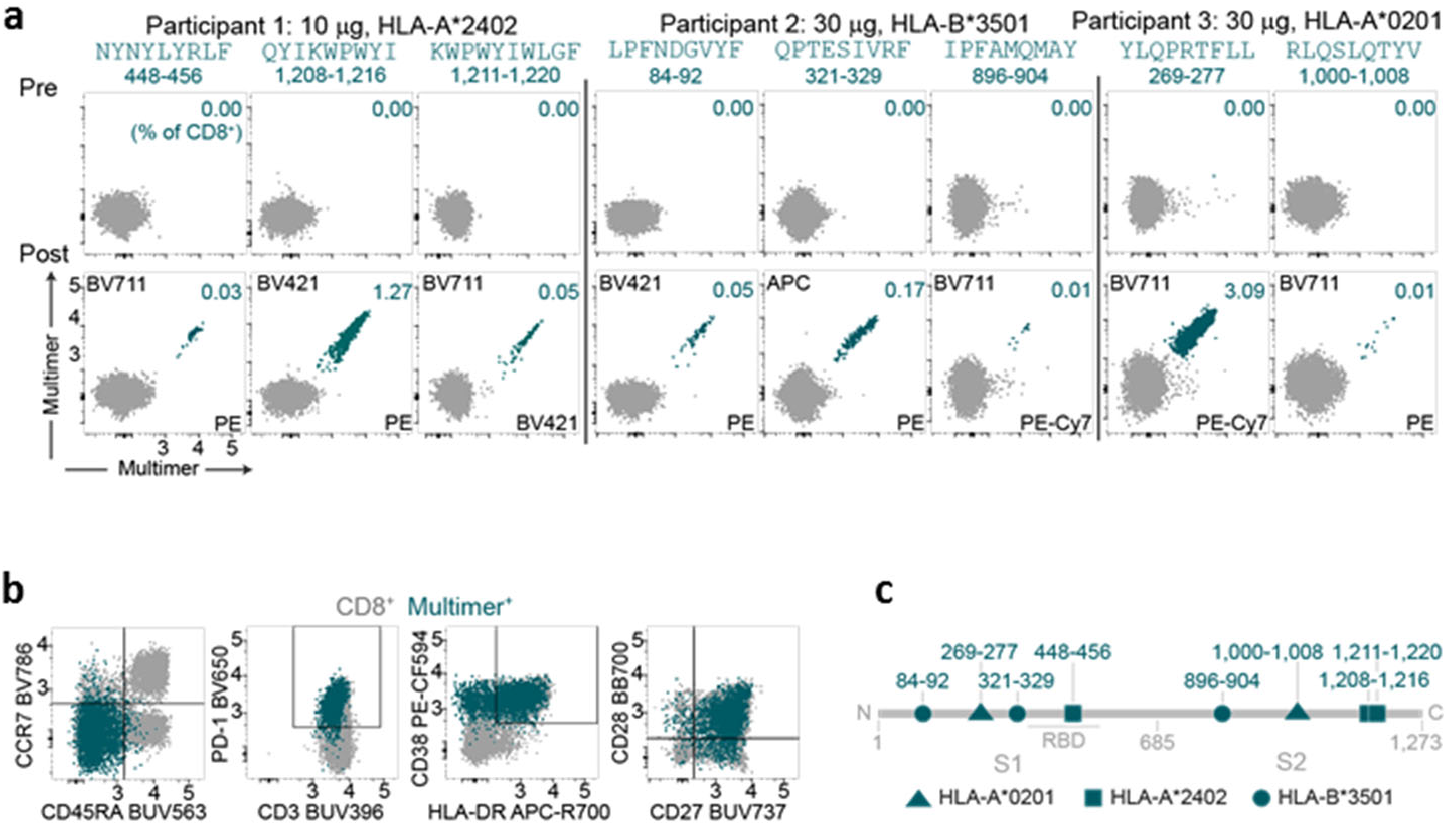
Characterization of BNT162b2-induced T cells on the single epitope level. PBMCs obtained on day 1 (pre-prime) and day 29 (7 days post-boost) of three vaccinated participants (dose cohorts 10 µg, *n*=1; 30 µg, *n*=2) were stained with individual pMHC class I multimer cocktails and analysed for T cell epitope specificity **(a)** and phenotype (**b**; example from participant 3; YLQPRTFLL) by flow cytometry (for gating strategy see Supplementary Fig. 2). Peptide sequences above dot plots indicate pMHC class I multimer epitope specificity, numbers above dot plots indicate the amino acids corresponding to the epitope within S. **c**, Localization of identified MHC class I-restricted epitopes within S.

Whereas the pMHC multimer approach probes a discrete subset of potential reactivities, bulk IFNγ^+^ CD8^+^ T cell responses against full S determined by ELISpot and ICS are considered to comprehensively capture the full poly-epitopic T cell response. However, comparison of both data sets indicated that a functional T cell assay may underestimate the true extent of the cellular immune response (Extended Data Fig. 5f).

Phenotyping of the identified pMHC multimer^+^ S antigen-experienced CD8^+^ T cell specificities revealed an early differentiated effector memory phenotype characterised by low expression of CCR7 and CD45RA and high expression of the costimulatory molecules CD28 and CD27. CD8^+^ T cells also expressed markers associated with cognate activation, such as CD38, HLA-DR and PD-1 (Fig. 4b).

## Discussion

Effectors of the adaptive immune system have complementary roles in the defense of viral infections. While neutralising antibodies are the first line of defense, CD8^+^ cytotoxic T lymphocytes (CTLs) contribute to virus clearance from intracellular compartments that are inaccessible to neutralising antibodies. Antigen-specific CD4^+^ T cells have immune orchestrating functions, including provision of cognate help to B cells and CD8^+^ T cells, support of memory generation, as well as indirect (*e*.*g*. via IFNγ) or direct (against MHC class II-expressing target cells) cytotoxic activity.

There is broad consensus reflected in the design of ongoing clinical trials that a COVID-19 vaccine should induce antibodies to SARS-CoV-2 S. However, it is not yet known if antibody responses will be sufficient for full and long-lasting protective immunity to SARS-CoV-2, and what the contribution of SARS-CoV-2-specific T cells may be.

Previous experience with the closely related first SARS-CoV suggests that T cells prevent severe forms of the disease^27^ and may be associated with long-term protection^28,29^. For the novel SARS-CoV-2, an understanding of mechanisms of immunity from studies of infected and convalescent individuals is only beginning to emerge. An increasing amount of data supports a role of T cell immune responses^30–32^. COVID19 patients with critical disease states were reported to lack S1-reactive CD4^+^ T cells^33^. Cases of asymptomatic virus exposure have been associated with cellular immune responses without seroconversion, indicating that SARS-CoV-2 specific T cells could be relevant in disease control even in the absence of neutralising antibodies^29^.

We report here that vaccination with BNT162b2 induces a coordinated immune response with SARS-CoV-2 S-specific neutralising antibodies, CD4^+^ T cells, CD8^+^ T cells, and immune-modulatory cytokines such as IFNγ.

All participants vaccinated with BNT162b2 mounted de novo S-specific CD4^+^ T cell responses and almost 92% of participants mounted CD8^+^ T cell responses, as detected with an ex vivo ELISpot assay. The magnitude of the T cell responses varied inter-individually and showed no clear dose dependency. Even with the lowest dose of 1 µg BNT162b2, most of the vaccinated participants demonstrated robust expansion of CD4^+^ and CD8^+^ T cells. T cell responses were directed against RBD, S1 and S2 regions of S, indicating immune recognition of multiple independent MHC I and II epitopes, which was one of the reasons to favour BNT162b2 over BNT162b1.

Expression of IFNγ and IL-2 but only low levels of IL-4 in BNT162b2-induced CD4^+^ T cells indicated a T_H_1 profile and the absence of a potentially deleterious T_H_2 immune response.

While all CD8^+^ T cell responses against the S1 subunit of S were de novo and not detected at baseline, pre-existing immune responses against the S2 subunit were identified in several individuals. The S1 fragment has less sequence similarity to the corresponding seasonal coronavirus sequences than the S2 fragment does, indicating that we may have detected pre-existing cross-reactive CD8^+^ T cells^34,35^.

pMHC multimer technology enabled the identification of S epitopes recognised by vaccine-induced CD8^+^ T cells as well as direct quantification of the respective epitope-specific T cells. The cumulative T cell frequencies in each participant exceeded the overall T cell response measured in ELISpot and ICS assays, indicating that those assays underestimate the true magnitude of the poly-epitopic response. Single peptide analyses are well known to yield higher T cell frequencies as compared to functional T cell assays that stimulate with peptide pools, with a multitude of immunogenic epitopes competing. A high proportion of induced CD8^+^ T cells were early differentiated effector memory cells. This favourable phenotype has the potential to respond rapidly, but has a limited capacity to produce IFNγ, and thus is less likely to be detected in functional T cell assays. Previous studies have identified epitopes in SARS-CoV-2 S against which infected individuals raise CD8^+^ T cells^36,37^. To our knowledge, this is the first report of epitopes recognised by COVID-19 vaccine-induced T cells. Of note, the immunodominant HLA-A*02:01 restricted peptide YLQPRTFLL identified in our study has previously been described in convalescent COVID-19 patients^36,37^.

In addition to providing new insights into T cell responses, this study reproduces our previous findings in the U.S.A trial^1^, and confirms a benign safety profile and robust induction of antibody responses, with the latter being followed up for a longer period, up to day 85 (63 days post-boost). Prime/boost vaccination with 10 to 30 µg of BNT162b2 elicited GMTs that, after an initial decline, remained stable up to day 85 in the range of, or higher than, GMTs in COVID-19 recovered individuals. BNT162b2 immune sera efficiently neutralised 19 pseudotyped viruses (18 of which enter cells using an S protein with a different RBD variant, and one of which uses the dominant S variant D614G), indicating the potential for broad BNT162b2-elicited protection against reported mutations^1,9,10^.

Limitations of our clinical study include the small sample size, the lack of representation of populations of interest (*e*.*g*. older adults, other ethnicities, immunocompromised individuals and pediatric populations), and limited availability of blood samples for a more in-depth T cell analysis.

Typically antigen-activated B and T cells go through proliferation, followed by rebound contraction with a gradual decline in numbers before entering a sustained memory phase^22,23^, and the short-term follow up presented in this paper does not allow for extrapolation of long-term durability of the immune responses. Whereas it is encouraging that BNT162b2 robustly activates antigen-specific humoral and as cellular immune effector systems, it is not clear whether this immune response pattern will protect from SARS-CoV-2 infection and prevent COVID-19. These questions will be addressed by the ongoing clinical program, which includes longer term follow-up of participants in the two ongoing phase 1/2 trials, a dedicated immune biomarker trial to further dissect the composite elements of the immune response, and the ongoing phase 2/3 study with efficacy endpoints.

## Data Availability

The data that support the findings of this study are available from the corresponding author upon reasonable request. Upon completion of this clinical trial, summary-level results will be made public and shared in line with data sharing guidelines.

## Acknowledgements

We thank Mikael Dolsten, Pfizer Chief Scientific Officer, for advice during drafting of the manuscript. We thank C. Anders, C. Anft, N. Beckmann, K. Bissinger, P. Cienskowski, K. Clarke, C. Ecker, A. Engelmann, M. Fierek, D. Harjanto, A. Heinen, M. Hossainzadeh, S. Jägle, L. Jeck, O. Kahl, D. Kallin, M. Knezovic, T. Kotur, M. Kretschmer, A. Kruithof, J. Mc Gee, B. Mehlhase, C. Müller, S. Murphy, L.-M. Schmid, K. Schmoldt, R. Schulz, L. Srinivasan, M. Vehreschild, T. Weisenburger and S. Wessel for technical support, project management and advice. We thank P. Koch for data management and analysis. We thank O. Kistner, S. Liebscher, J. Loschko and K. Swanson for expert advice. We thank Judith Absalon for manuscript advice. We thank the CRS Team (Mannheim and Berlin) for study conduct: S. Armani, M. Berse, M. Casjens, B. Ehrlich, F. Seitz, M. Streckebein. We thank W. Kalina, I. Scully, and the Pfizer Vaccines Clinical Assays Team and the Pfizer Aviation Team for technical and logistical support of serology analyses. We thank GISAID Nucleotide database for sharing of SARS-CoV-2 complete genome sequences.

## Author Contributions

U.S. conceived and conceptualised the work and strategy supported by Ö.T. Experiments were planned or supervised by N.B., E.D., C. F.-G, C.A.K., U.L., A.M., J.Q., P.-Y.S., A.U. and I.V.. A.B., N.B., D.C, M.C., C. F.-G, K.P., J.Q., A.U. and P.-Y.S. performed experiments. D.B., S. Brachtendorf, E.D., P.R.D., J.G., K.U.J., A.-K.E., P.K., M.T., L.M.K., M.-C.K., V.L., A.M., J.Q., J.S., N.B., A.U., I.V. and M.V. analysed data. A.P. prioritised epitopes for multimer assay. J.Z.D. supervised manufacturing and delivery of peptides for multimer assay. D.M. planned and supervised dashboards for analysis of clinical trial data. R.H. was responsible for data normalization and adaption. C.B, L.H. and C.R. were responsible for biomarker and R&D program management. G.B., K.K., A.J.M., J.R. and G.T.S. optimised mRNA characteristics. A.K-B., S. Baumann, A.S., D.L., M.B., S. Bolte, and T.P. coordinated operational conduct of the clinical trial. J.L.P. advised on the trial. U.S., Ö.T., supported by M.B., N.B., E.D., P.R.D., K.U.J., L.M.K., A.M., A.U., I.V. and M.V., interpreted data and wrote the manuscript. All authors supported the review of the manuscript.

## Additional Information

Supplementary Information is available for this paper.

Correspondence and requests for materials should be addressed to Ugur Sahin.

## Materials and Methods

### Clinical trial design

Study BNT162-01 (NCT04380701) is an ongoing, umbrella-type first-in-human, phase 1/2, open-label, dose-ranging clinical trial to assesses the safety, tolerability, and immunogenicity of ascending dose levels of various intramuscularly administered BNT162 mRNA vaccine candidates in healthy men and non-pregnant women 18 to 55 years (amended to add 56-85 years) of age. The principle endpoints of the study are safety and immunogenicity. Key exclusion criteria included previous clinical or microbiological diagnosis of COVID-19; receipt of medications to prevent COVID-19; previous vaccination with any coronavirus vaccine; a positive serological test for SARS-CoV-2 IgM and/or IgG; and a SARS-CoV-2 nucleic acid amplification test (NAAT)-positive nasal swab; increased risk for severe COVID-19; and immunocompromised individuals.

The presented data are from the BNT162b2-immunised healthy adults 19 to 55 years of age exposed to dose levels 1, 10, 20 or 30 μg. The data are based on a preliminary analysis (data extraction date of 23 October 2020 for safety and antibody analysis, 16 October 2020 and 24 November 2020 for T cell analysis [intracellular cytokine staining and ELISpot, respectively]) and are focused on analysis of vaccine-induced immunogenicity descriptively summarised at the various time points, and on reactogenicity. All participants with data available were included in the immunogenicity analyses. This part of the study was performed at one site in Germany with 12 healthy participants per dose level in a dose-escalation/de-escalation design. Sentinel dosing was performed in each dose-escalation cohort. Progression in that cohort and dose escalation required data review by a safety review committee. Participants received a BNT162b2 priming dose on day 1, and a booster dose on day 22±2 (on day 28 for one participant from the 10 µg dose cohort). Serum for antibody assays was obtained on day 1 (pre-prime), 8±1 (post-prime), 22±2 (pre-boost), 29±3, 43±4 and 50±4 (post-boost). PBMCs for T cell studies were obtained on day 1 (pre-prime) and 29±3 (post-boost) (Extended Data Fig. 1). Follow-up of participants is ongoing and includes assessment of antibody and T cell responses at later time points. Reactogenicity was assessed by patient diary. Two participants discontinued prior to the booster dose due to study drug-unrelated withdrawal (participant in the 1 µg dose cohort) and an adverse event (participant of the 10 µg dose cohort) (upper respiratory syndrome), respectively. The trial was carried out in Germany in accordance with the Declaration of Helsinki and Good Clinical Practice Guidelines and with approval by an independent ethics committee (Ethik-Kommission of the Landesärztekammer Baden-Württemberg, Stuttgart, Germany) and the competent regulatory authority (Paul-Ehrlich Institute, Langen, Germany). All participants provided written informed consent.

### Manufacturing of RNA

BNT162b2 incorporates a Good Manufacturing Practice (GMP)-grade mRNA drug substance that encodes the trimerised SARS-CoV-2 S glycoprotein RBD antigen. The RNA is generated from a DNA template by *in vitro* transcription in the presence of 1-methylpseudouridine-5’-triphosphate (m1ΨTP; Thermo Fisher Scientific) instead of uridine-5’-triphosphate (UTP). Capping is performed co-transcriptionally using a trinucleotide cap 1 analogue ((m_2_^7,3’-O^)Gppp(m^2’-O^)ApG; TriLink). The antigen-encoding RNA contains sequence elements that increase RNA stability and translation efficiency in human dendritic cells^11,12^. The mRNA is formulated with lipids to obtain the RNA-LNP drug product. The vaccine was transported and supplied as a buffered-liquid solution for IM injection and was stored at −80 °C.

### Proteins and peptides

Two pools of 15-mer peptides overlapping by 11 amino acids (aa) and together covering the whole sequence of wild-type SARS-CoV-2 S (S pool 1 featuring aa 1-643, S pool 2 featuring aa 633-1273) and one pool covering the SARS-CoV-2 RBD (aa 327-528) with the signal peptide of S (aa 1-16) fused to its N-terminus were used for ex vivo stimulation of PBMCs for flow cytometry and IFNγ ELISpot. CEF (CMV, EBV, influenza virus; human leukocyte antigen [HLA] class I epitope peptide pool) and CEFT (CMV, EBV, influenza virus, tetanus toxoid; HLA class II epitope peptide pool) were used as controls for general T cell reactivity and to benchmark the magnitude of memory T cell responses. All the above peptides were obtained from JPT Peptide Technologies. The 8-12 amino acid long peptides used in the easYmer assays were produced at BioNTech US.

### Human convalescent serum and PBMC panel

Human SARS-CoV-2 infection/COVID-19 convalescent sera (*n*=38) were drawn from donors 18-83 years of age at least 14 days after PCR-confirmed diagnosis and at a time when the participants were asymptomatic. The mean age of the donors was 45 years. Neutralising GMTs in subgroups of the donors were as follows: symptomatic infections, 90 (*n*=35); asymptomatic infections, 156 (*n*=3); hospitalized, 618 (*n*=1). Sera were obtained from Sanguine Biosciences (Sherman Oaks, CA), the MT Group (Van Nuys, CA) and Pfizer Occupational Health and Wellness (Pearl River, NY). Human SARS-CoV-2 infection/COVID-19 convalescent PBMC samples (*n*=18) were collected from donors 22-79 years of age 30-62 days after PCR-confirmed diagnosis, when donors were asymptomatic. PBMC donors had asymptomatic or mild infections (*n*=16, clinical score 1 and 2) or had been hospitalized (*n*=2, clinical score 4 and 5). Blood samples were obtained from the Frankfurt University Hospital.

### Cell culture and primary cell isolation

Vero cells (American Type Culture Collection [ATCC] CCL-81) and Vero E6 cells (ATCC CRL-1586) were cultured in Dulbecco’s modified Eagle’s medium (DMEM) with GlutaMAX™ (Gibco) supplemented with 10% fetal bovine serum (FBS) (Sigma-Aldrich). Cell lines were tested for mycoplasma contamination after receipt and before expansion and cryopreservation. PBMCs were isolated by Ficoll-Paque™ PLUS (Cytiva) density gradient centrifugation and cryopreserved prior to analysis.

### S1- and RBD-binding IgG assay

Recombinant SARS-CoV-2 S1 or RBD containing a C-terminal Avitag™ (Acro Biosystems) were bound to streptavidin-coated Luminex microspheres. Heat-inactivated participant sera were diluted 1:500, 1:5,000, and 1:50,000. Following an overnight incubation at 2-8 °C while shaking, plates were washed in a solution containing 0.05% Tween-20. A secondary fluorescently labelled goat anti-human polyclonal antibody (Jackson Labs) was added for 90 minutes at room temperature while shaking, before plates were washed once more in a solution containing 0.05% Tween-20. Data were captured as median fluorescent intensities (MFIs) using a Bioplex200 system (Bio-Rad) and converted to U/mL antibody concentrations using a reference standard curve with arbitrarily assigned concentrations of 100 U/mL and accounting for the serum dilution factor. The reference standard was composed of a pool of five convalescent serum samples obtained >14 days after COVID-19 PCR diagnosis and was diluted sequentially in antibody-depleted human serum. Three dilutions were used to increase the likelihood that at least one result for any sample would fall within the useable range of the standard curve. Assay results were reported in U/mL of IgG. The final assay results were expressed as the geometric mean concentration of all sample dilutions that produced a valid assay result within the assay range.

### SARS-CoV-2 neutralisation assay

The neutralisation assay used a previously described strain of SARS-CoV-2 (USA_WA1/2020) that had been rescued by reverse genetics and engineered by the insertion of an mNeonGreen (mNG) gene into open reading frame 7 of the viral genome^38^. This reporter virus generates similar plaque morphologies and indistinguishable growth curves from wild-type virus. Viral master stocks (2 × 10^7^ PFU/mL) were grown in Vero E6 cells as previously described^38^. With patient convalescent sera, the fluorescent neutralisation assay produced comparable results to the conventional plaque reduction neutralisation assay^39^. Serial dilutions of heat-inactivated sera were incubated with the reporter virus (2 × 10^4^ PFU per well to yield a 10-30% infection rate of the Vero CCL81 monolayer) for 1 hour at 37 °C before inoculating Vero CCL81 cell monolayers (targeted to have 8,000 to 15,000 cells in a central field of each well at the time of seeding, 24 hours before infection) in 96-well plates to allow accurate quantification of infected cells. Total cell counts per well were enumerated by nuclear stain (Hoechst 33342) and fluorescent virally infected foci were detected 16-24 hours after inoculation with a Cytation 7 Cell Imaging Multi-Mode Reader (BioTek) with Gen5 Image Prime version 3.09. Titers were calculated in GraphPad Prism version 8.4.2 by generating a 4-parameter (4PL) logistical fit of the percent neutralisation at each serial serum dilution. The 50% neutralisation titre (VNT_50_) was reported as the interpolated reciprocal of the dilution yielding a 50% reduction in fluorescent viral foci.

### VSV-SARS-CoV-2 S variant pseudovirus neutralisation assay

Vesicular stomatitis virus (VSV)-SARS-CoV-2-S pseudoparticle generation and neutralisation assays were performed as previously described^24^. Briefly, human codon optimized SARS-CoV-2 S (GenBank: MN908947.3) was synthesised (Genscript) and cloned into an expression plasmid. SARS-CoV-2 complete genome sequences were downloaded from GISAID nucleotide database (https://www.gisaid.org). Sequences were curated, and genetic diversity of the S-encoding gene was assessed across high quality genome sequences using custom pipelines. Amino acid substitutions were cloned into the S expression plasmid using site-directed mutagenesis. HEK293T cells (ATCC CRL-3216) were seeded (culture medium: DMEM high glucose [Life Technologies] supplemented with 10% heat-inactivated FBS (Life Technologies) and penicillin/streptomycin/L-glutamine [Life Technologies]) and transfected the following day with S expression plasmid using Lipofectamine LTX (Life Technologies) following the manufacturer’s protocol. At 24 hours post-transfection at 37 °C, cells were infected with the VSVΔG:mNeon/VSV-G diluted in Opti-MEM (Life Technologies) at a multiplicity of infection of 1. Cells were incubated 1 hour at 37 °C, washed to remove residual input virus and overlaid with infection medium (DMEM high glucose supplemented with 0.7% Low IgG bovine serum albumin [BSA, Sigma], sodium pyruvate [Life Technologies] and 0.5% Gentamicin [Life Technologies]). After 24 hours at 37 °C, the medium containing VSV-SARS-CoV-2-S pseudoparticles was collected, centrifuged at 3000 x g for 5 minutes to clarify and stored at −80 °C until further use.

For pseudovirus neutralisation assays, Vero cells (ATCC CCL-81) were seeded in 96-well plates in culture medium and allowed to reach approximately 85% confluence before use in the assay (24 hours later). Sera were serially diluted 1:2 in infection medium starting with a 1:300 dilution. VSV-SARS-CoV-2-S pseudoparticles were diluted 1:1 in infection medium for a fluorescent focus unit (ffu) count in the assay of ∼1000. Serum dilutions were mixed 1:1 with pseudoparticles for 30 minutes at room temperature prior to addition to Vero cells and incubation at 37 °C for 24 hours. Supernatants were removed and replaced with PBS (Gibco), and fluorescent foci were quantified using the SpectraMax i3 plate reader with MiniMax imaging cytometer (Molecular Devices). Neutralisation titers were calculated in GraphPad Prism version 8.4.2 by generating a 4-parameter logistical (4PL) fit of the percent neutralisation at each serial serum dilution. The 50% pseudovirus neutralisation titre (pVNT_50_) was reported as the interpolated reciprocal of the dilution yielding a 50% reduction in fluorescent viral foci.

### IFNγ ELISpot

IFNγ ELISpot analysis was performed ex vivo (without further in vitro culturing for expansion) using PBMCs depleted of CD4^+^ and enriched for CD8^+^ T cells (CD8^+^ effectors) or depleted of CD8^+^ and enriched for CD4^+^ T cells (CD4^+^ effectors). Tests were performed in duplicate and with a positive control (anti-CD3 monoclonal antibody CD3-2 [1:1,000; Mabtech]). Multiscreen filter plates (Merck Millipore) pre-coated with IFNγ-specific antibodies (ELISpotPro kit, Mabtech) were washed with PBS and blocked with X-VIVO 15 medium (Lonza) containing 2% human serum albumin (CSL-Behring) for 1-5 hours. Per well, 3.3 × 10^5^ effector cells were stimulated for 16-20 hours with three overlapping peptide pools representing different portions of the wild-type sequence of SARS-CoV-2 S (N-terminal pools S pool 1 [aa 1-643] and RBD [aa1-16 fused to aa 327-528], and the C-terminal S pool 2 [aa 633-1273]). Bound IFNγ was visualised using a secondary antibody directly conjugated with alkaline phosphatase followed by incubation with 5-bromo-4-chloro-3′-indolyl phosphate (BCIP)/ nitro blue tetrazolium (NBT) substrate (ELISpotPro kit, Mabtech). Plates were scanned using an AID Classic Robot ELISPOT Reader and analysed by AID ELISPOT 7.0 software (AID Autoimmun Diagnostika). Spot counts were displayed as mean values of each duplicate. T cell responses stimulated by peptides were compared to effectors incubated with medium only as a negative control using an in-house ELISpot data analysis tool (EDA), based on two statistical tests (distribution-free resampling) according to Moodie et al.^40,41^, to provide sensitivity while maintaining control over false positives.

To account for varying sample quality reflected in the number of spots in response to anti-CD3 antibody stimulation, a normalisation method was applied, enabling direct comparison of spot counts and strength of response between individuals. This dependency was modelled in a log-linear fashion with a Bayesian model including a noise component (unpublished). For a robust normalisation, each normalisation was sampled 10,000 times from the model and the median taken as normalised spot count value. Likelihood of the model: log *λ*_*E*_*= α* log*λ*_*P*_ *+* log*β*_*j*_ + *σ ε*, where *λ*_*E*_ is the normalized spot count of the sample; *α* is a stable factor (normally distributed) common among all positive controls *λ*_*P*_; *β*_*j*_ is a sample *j* specific component (normally distributed); and *σ ε* is the noise component, of which *σ* is Cauchy distributed, and *ε* is Student’s-t distributed. *β*_*j*_ ensures that each sample is treated as a different batch.

### Flow cytometry

Cytokine-producing T cells were identified by intracellular cytokine staining. PBMCs thawed and rested for 4 hours in OpTmizer medium supplemented with 2 µg/mL DNase I (Roche), were restimulated with different portions of the wild-type sequence of SARS-CoV-2 S in peptide pools described in the ELISpot section (2 µg/mL/peptide; JPT Peptide Technologies) in the presence of GolgiPlug (BD) for 18 hours at 37 °C. Controls were treated with DMSO-containing medium. Cells were stained for viability and surface markers (CD3 BV421, 1:250; CD4 BV480, 1:50; CD8 BB515, 1:100; all BD Biosciences) in flow buffer (DPBS [Gibco] supplemented with 2% FBS [Biochrom], 2 mM ethylenediaminetetraacetic acid [EDTA; Sigma-Aldrich]) for 20 minutes at 4 °C. Afterwards, samples were fixed and permeabilised using the Cytofix/Cytoperm kit according to manufacturer’s instructions (BD Biosciences). Intracellular staining (CD3 BV421, 1:250; CD4 BV480, 1:50; CD8 BB515, 1:100; IFNγ PE-Cy7, 1:50 [for HCS]; IFNγ BB700, 1:250 [for participants]; IL-2 PE, 1:10; IL-4 APC, 1:500; all BD Biosciences) was performed in Perm/Wash buffer for 30 minutes at 4 °C. Samples were acquired on a fluorescence-activated cell sorter (FACS) VERSE instrument (BD Biosciences) and analysed with FlowJo software version 10.6.2 (FlowJo LLC, BD Biosciences). S- and RBD-specific cytokine production was corrected for background by subtraction of values obtained with dimethyl sulfoxide (DMSO)-containing medium. Negative values were set to zero. Cytokine production in Figure 4b was calculated by summing up the fractions of all CD4^+^ T cells positive for either IFNγ, IL-2 or IL-4, setting this sum to 100% and calculating the fraction of each specific cytokine-producing subset thereof. Pseudocolor plot axes are in log10 scale.

### Peptide/MHC multimer staining

In order to select MHC-class I epitopes for multimer analysis, a mass spectrometry-based binding and presentation predictor^42,43^ was applied to 8-12 amino acid long peptide sequences from the Spike glycoprotein derived from the GenBank reference sequence for SARS-CoV-2 (accession: NC_045512.2, https://www.ncbi.nlm.nih.gov/nuccore/NC_045512) and paired with 18 MHC-class-I alleles with >5% frequency in the European population. Top predicted epitopes were identified by setting thresholds to the binding percent-rank (≤1%) and presentation scores (≥10^−2.2^). Peptides were manufactured at >90% purity. pMHC complexes were refolded with the easYmer technology (easYmer® kit, ImmuneAware Aps), and complex formation was validated in a bead-based flow cytometry assay according to the manufacturer’s instructions^44,45^. Combinatorial labeling was used for dissecting the antigen specificity of T cells utilizing two-color combinations of five different fluorescent labels to enable detection of up to ten different T cell populations per sample^46^. For tetramerisation, streptavidin (SA)-fluorochrome conjugates were added: SA BV421, SA BV711, SA PE, SA PE-Cy7, SA APC (all BD Biosciences). For three BNT162b2 vaccinated participants, individualized pMHC multimer staining cocktails contained up to ten pMHC complexes, with each pMHC complex encoded by a unique two-color combination. PBMCs (2×10^6^) were stained ex vivo for 20 minutes at room temperature with each pMHC multimer cocktail at a final concentration of 4 nM in Brilliant Staining Buffer Plus (BSB Plus [BD Horizon™]). Surface and viability staining was carried out in flow buffer (DPBS [Gibco] with 2% FBS [Biochrom], 2 mM EDTA [Sigma-Aldrich]) supplemented with BSB Plus for 30 minutes at 4 °C (CD3 BUV395, 1:50; CD45RA BUV563, 1:200; CD27 BUV737, 1:200; CD8 BV480, 1:200; CD279 BV650, 1:20; CD197 BV786, 1:15; CD4 BB515, 1:50; CD28 BB700, 1:100; CD38 PE-CF594, 1:600; HLA-DR APC-R700, 1:150; all BD Biosciences; DUMP channel: CD14 APC-eFluor780, 1:100; CD16 APC-eFluor780, 1:100; CD19 APC-eFluor780, 1:100; fixable viability dye eFluor780, 1:1,667; all ThermoFisher Scientific). Cells were fixed for 15 minutes at 4 °C in 1x Stabilization Fixative (BD), acquired on a FACSymphony™ A3 flow cytometer (BD Biosciences) and analysed with FlowJo software version 10.6.2 (FlowJo LLC, BD Biosciences). CD8^+^ T cell reactivities were considered positive, when a clustered population was observed that was labelled with only two pMHC multimer colors.

### Statistical analysis

The sample size for the reported part of the study was not based on statistical hypothesis testing. All participants with data available were included in the safety and immunogenicity analyses. The statistical method of aggregation used for the analysis of antibody concentrations and titers is the geometric mean and the corresponding 95% CI. Employing the geometric mean accounts for non-normal distribution of antibody concentrations and titers spanning several orders of magnitude. Spearman correlation was used to evaluate the monotonic relationship between non-normally distributed data sets.

All statistical analyses were performed using GraphPad Prism software version 8.4.2.

## Extended Data Figures and Tables

**Extended Data Figure 1.**
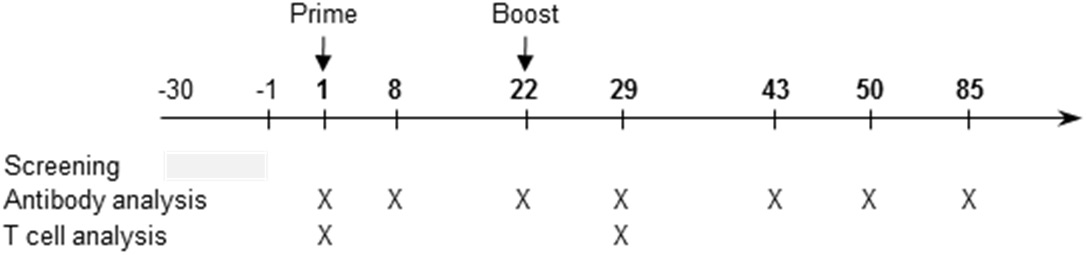
Schedule of vaccination and assessment. Study participants received a priming immunisation with BNT162b2 on day 1, and a booster immunisation on day 22±2. Serum was obtained on days 1 (pre-prime), 8±1 (post-prime), 22±2 (pre-boost), 29±3, 43±4, 50±4 and 85±7 (post-boost). PBMCs were obtained on days 1 (pre-prime) and 29±3 (post-boost).

**Extended Data Figure 2.**
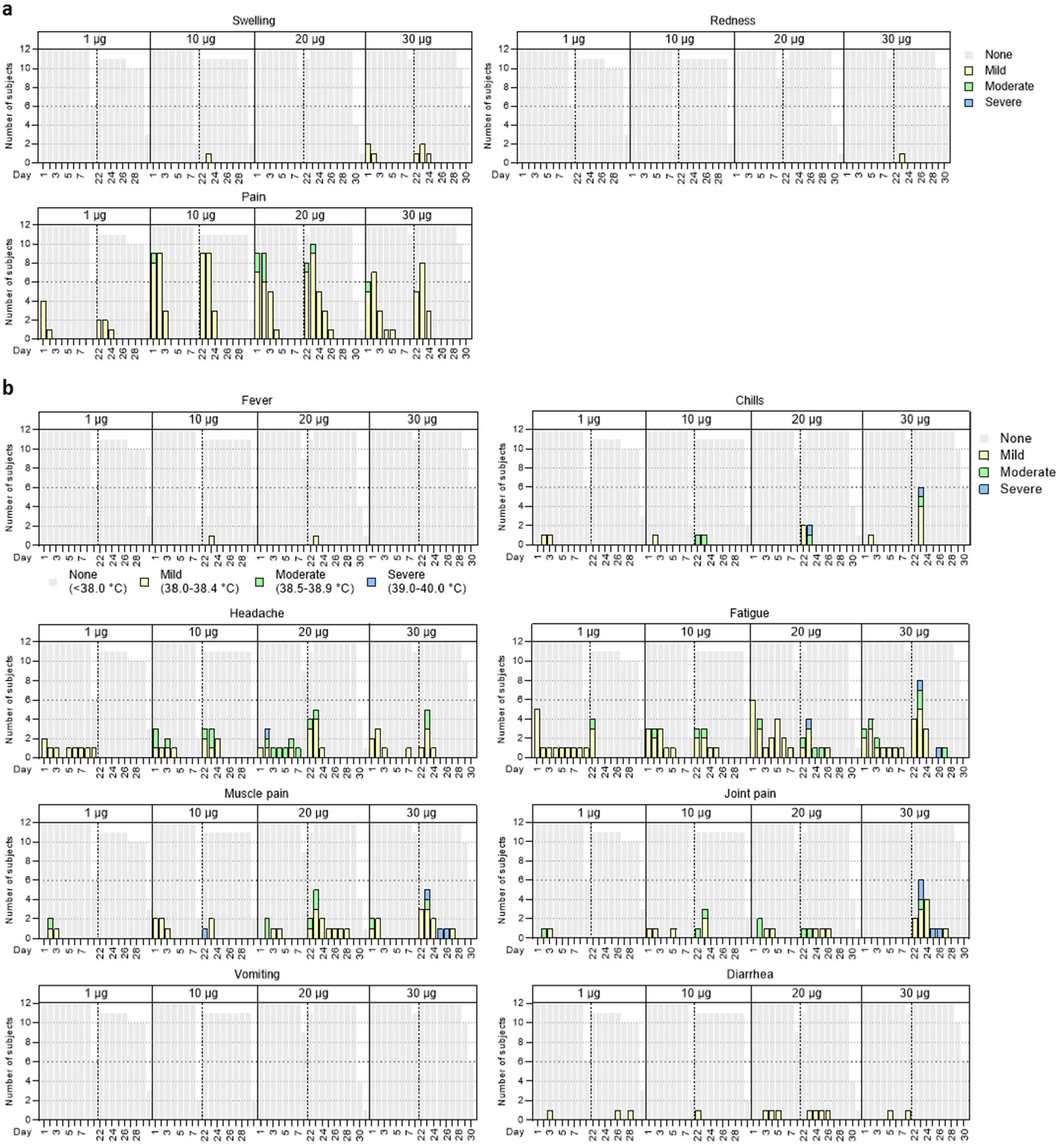
Solicited adverse events. Number of participants with local (**a**) or systemic solicited adverse events (AE) (**b**). Participants were immunised with BNT162b2 on days 1 and 22 (prime: *n*=12 per dose cohort; boost: 1, 10 µg, *n*=11); discontinuation of participants due to non-vaccine related reasons). Grey shading indicates number of participants at each time point. As per protocol, AEs were recorded up to 7 days after each immunisation (days 1-7 and 22-28) to determine reactogenicity; for some participants, 1-3 additional days of follow-up were available. Grading of adverse events was performed according to US Food and Drug Administration (FDA) recommendations^47^.

**Extended Data Figure 3.**
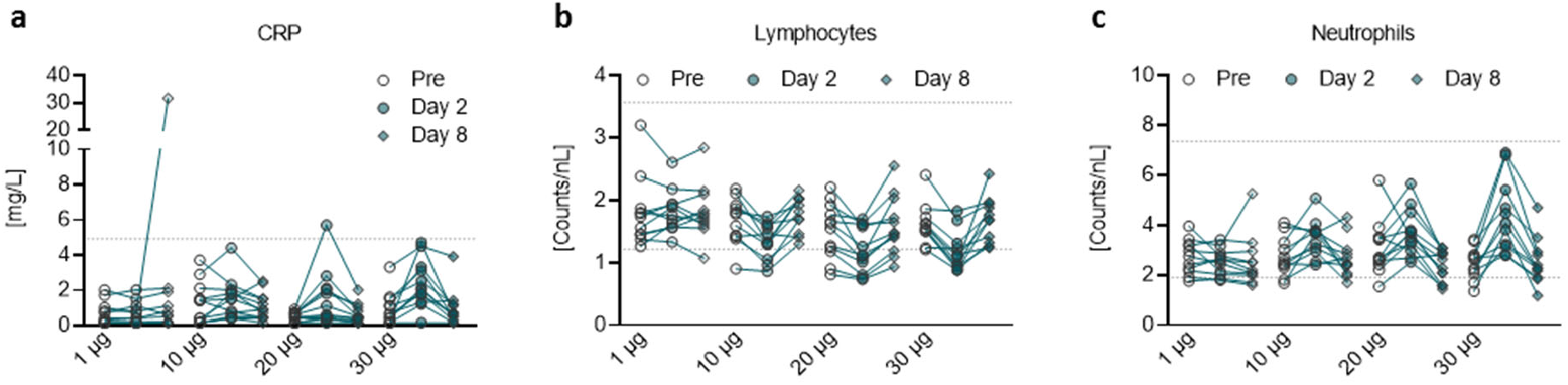
Pharmacodynamic markers. Participants were immunised with BNT162b2 on days 1 and 22 (*n*=12 per dose cohort). One participant in the 1 µg dose cohort (outlier on day 8 in [a] and highest data set in [b]) presented with a non-treatment related gastroenteritis on days 6 to 8. **a**, Kinetics of C-reactive protein (CRP) level. **b**, Kinetics of lymphocyte counts. **c**, Kinetics of neutrophil counts. Dotted lines indicate upper and lower limit of reference range. For values below the lower limit of quantification (LLOQ) = 0.3, LLOQ/2 values were plotted (**a**).

**Extended Data Figure 4.**
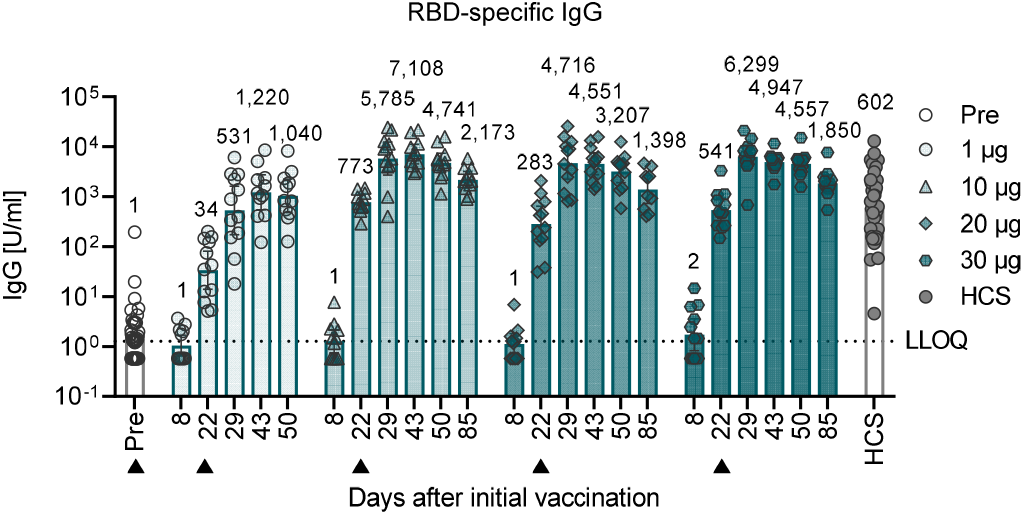
BNT162b2-induced RBD-specific IgG concentrations. Recombinant RBD-binding IgG geometric mean concentration in participants immunised with BNT162b2 on days 1 and 22 (*n*=12 per dose cohort; from day 22 onwards *n*=11 for the 1 µg and 10 µg dose cohorts). Vaccination schedule and serum sampling are described in Extended Data Fig. 1. Arrowheads indicate days of vaccination. Pre-dose responses across all dose levels were combined. COVID-19 convalescent samples (HCS, *n*=38) were obtained at least 14 days after PCR-confirmed diagnosis and at a time when the donors were no longer symptomatic. Each serum was tested in duplicate and geometric mean concentrations plotted. For values below LLOQ = 1.15, LLOQ/2 values were plotted. Group geometric mean concentrations (values above bars) with 95% confidence interval.

**Extended Data Figure 5.**
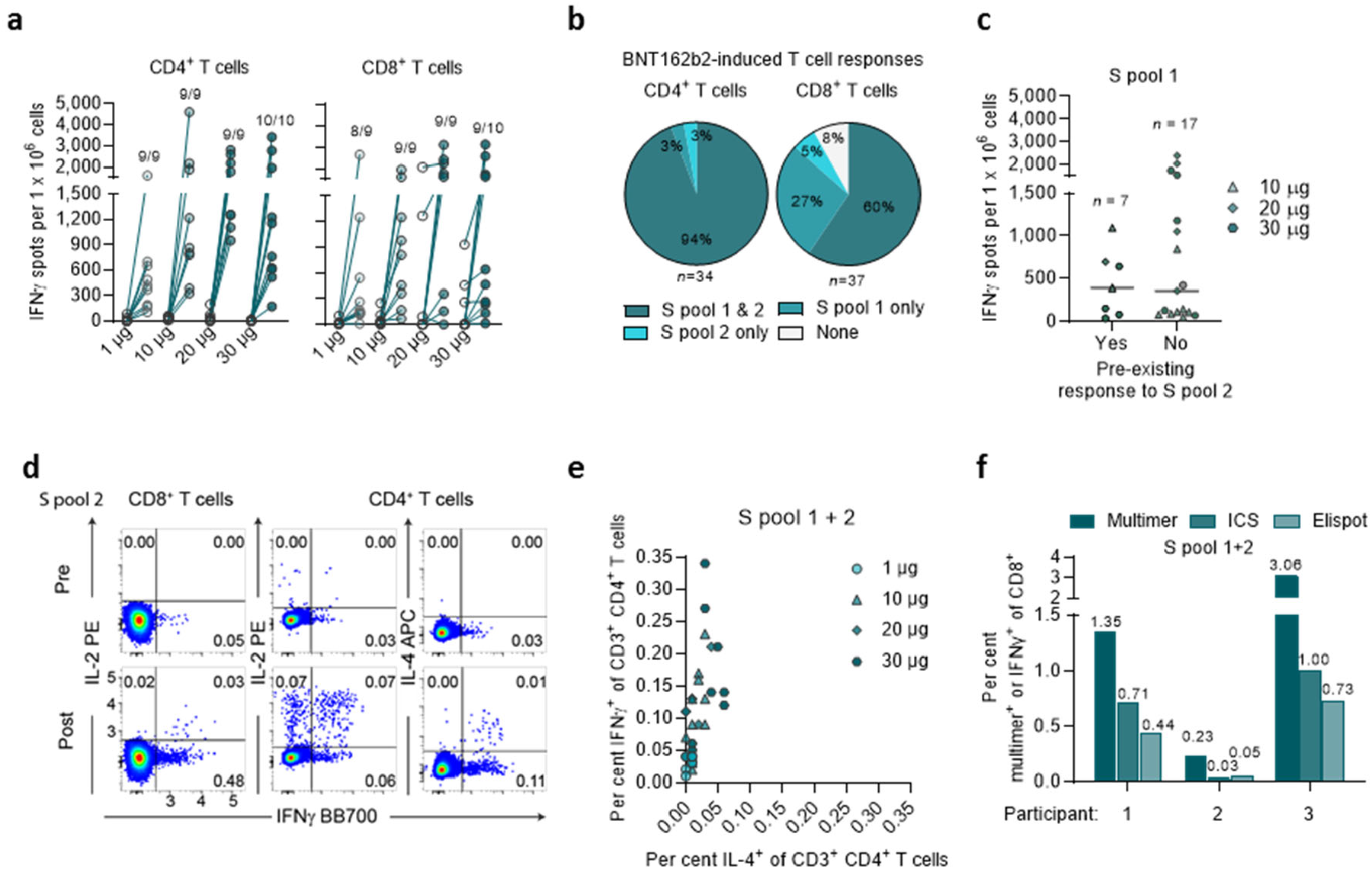
BNT162b2-induced S-specific CD8^+^ and CD4^+^ T cells. CD4^+^ or CD8^+^ T cell effector-enriched fractions of immunised participants derived from PBMCs obtained on day 1 (pre-prime) and day 29 (7 days post-boost) (1, 10 and 20 µg dose cohorts, *n*=9 each; 30 µg dose cohort, *n*=10) were stimulated overnight with two overlapping peptide pools covering the wild-type SARS-CoV-2 S (S pool 1 and S pool 2) for assessment in direct ex vivo IFNγ ELISpot (**a-c**). Each dot represents the normalised mean spot count from duplicate wells for one study participant, after subtraction of the medium-only control. T cell responses against S pool 1 and S pool 2 per participant were combined. Spot count data from two participants from the 20 µg dose cohort could not be normalised and are not plotted. PBMCs from vaccinated participants on day 29 (7 days post-boost) (dose cohorts 1 µg, *n*=7; 10 and 30 µg, *n*=10; 20 µg, *n*=9) were stimulated as described above and analysed by flow cytometry (**d, e**). **a**, S-specific CD4^+^ and CD8^+^ T cell responses for each dose cohort. Number of participants with detectable T cell response on day 29 over the total number of tested participants per dose cohort is provided. **b**, Mapping of vaccine-induced responses of participants with evaluable baseline data (*n*=34 for CD4^+^ and *n*=37 for CD8^+^ T cell responses) to different portions of S. De novo induced or amplified responses are classified as BNT162b2-induced response; no responses or pre-existing responses that were not amplified by the vaccinations are classified as no vaccine response (none). **c**, Response strength to S pool 1 in individuals with or without a pre-existing response to S pool 2. Data from the 1 µg dose cohort are excluded, as no baseline response to S pool 2 was present in this dose cohort. Horizontal bars represent median of each group. **d**, Examples of pseudocolor flow cytometry plots of cytokine-producing CD4^+^ and CD8^+^ T cells from a participant prime/boost vaccinated with 30 µg BNT162b2. **e**, Frequency of vaccine-induced, S-specific IFNγ^+^ CD4^+^ T cells vs. IL4^+^ CD4^+^ T cells. ICS stimulation was performed using a peptide mixture of S pool 1 and S pool 2. Each data point represents one study participant (1 µg dose cohort, n=8; 20 µg dose cohort, n=8; 10 and 30 µg, n=10 each). One participant from the 20 µg dose cohort with a strong pre-existing CD4^+^ T cell response to S pool 2 was excluded. **f**, Antigen-specific CD8^+^ T cell frequencies determined by pMHC class I multimer staining (% multimer^+^ of CD8^+^), ICS and ELISpot (% IFNγ^+^ of CD8^+^) for the three participants analysed in Figure 4. Signals for S pool 1 and S pool 2 were merged.

**Extended Data Figure 6.**
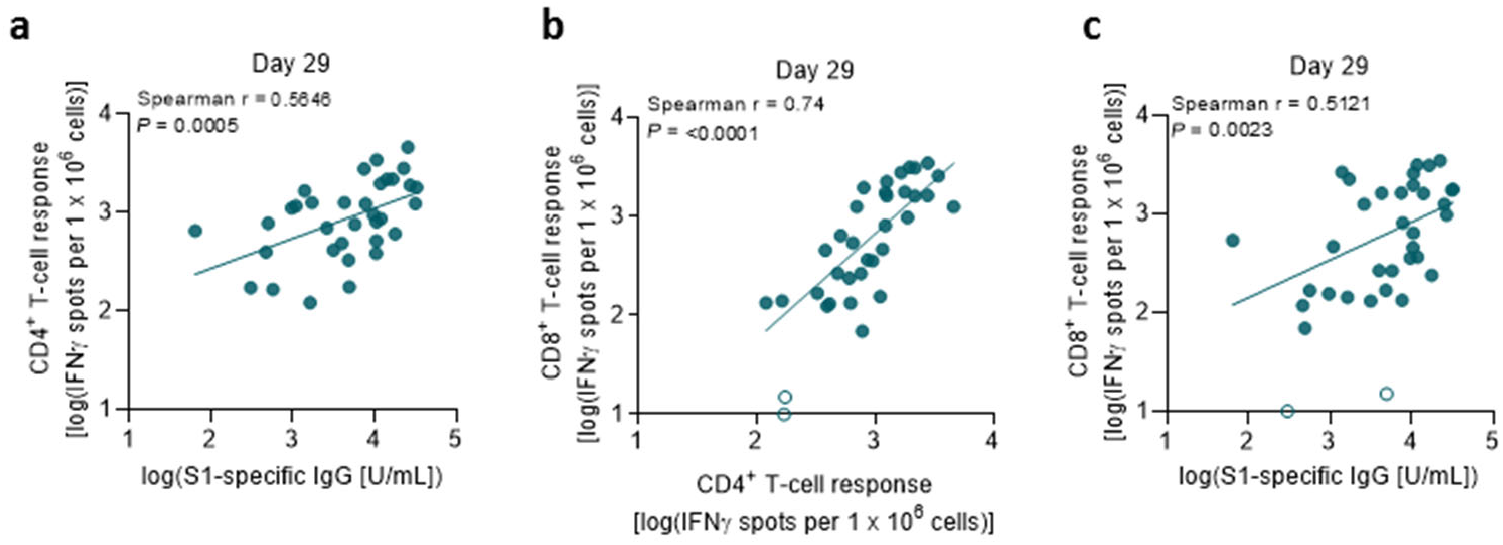
Correlation of antibody and T cell responses. Data are plotted for all prime/boost vaccinated participants (dose cohorts 1, 10, 20 and 30 µg) from day 29, with data points for participants with no detectable T cell response (open circles; **b, c**) excluded from correlation analysis. S1-specific IgG responses as in Fig. 1a, S-specific T cell responses as in Extended Data Fig. 5a (*n*=37). Nonparametric Spearman correlation. **a**, Correlation of S1-specific IgG responses with S-specific CD4^+^ T cell responses. **b**, Correlation of S-specific CD4^+^ with CD8^+^ T cell responses. **c**, Correlation of S1-specific IgG responses with S-specific CD8^+^ T cell responses.

**Extended Data Table 1.** Demographic characteristics.

| Cohort |  | 1 µg<br>(N=12)<br>n (%) | 10 µg<br>(N=12)<br>n (%) | 20 µg<br>(N=12)<br>n (%) | 30 µg<br>(N=12)<br>n (%) | Total<br>(N=48)<br>n (%) |
| --- | --- | --- | --- | --- | --- | --- |
| Sex | Male | 7 (58.3) | 4 (33.3) | 2 (16.7) | 8 (66.7) | 21 (43.8) |
|  | Female | 5 (41.7) | 8 (66.7) | 10 (83.3) | 4 (33.3) | 27 (56.2) |
| Race | Caucasian | 12 (100) | 12 (100) | 12 (100) | 12 (100) | 48 (100) |
|  | African American | 0 | 0 | 0 | 0 | 0 |
|  | Asian | 0 | 0 | 0 | 0 | 0 |
| Age at vaccination<br>(years) | Mean (SD) | 36.1 (10.09) | 34.8 (10.41) | 42.3 (9.86) | 46.7 (6.41) | 39.9 (10.26) |
|  | Median | 37.0 | 35.5 | 41.5 | 47.0 | 41.0 |
|  | Min, Max | 21, 53 | 19, 51 | 29, 55 | 35, 55 | 19, 55 |

N, number of participants in the specified group. This value is the denominator for the
percentage calculations. n, number of participants with the specified characteristics.

**Extended Data Table 2.** Participant disposition and analysis sets. Twelve participants per dose cohort received the priming and the booster dose except for two participants, who discontinued prior the booster dose due to a study drug-unrelated withdrawal by the participant (1 µg dose) and an adverse event (10 µg; upper respiratory syndrome), respectively. Safety analysis: Number of participants for whom 7 days of reactogenicity follow-up after both doses was evaluable at data cut-off. Antibody analysis: Numbers of participants for whom virus neutralisation assays and S1- and RBD-binding IgG antibody assays were performed. T cell analysis: Numbers of participants for whom PBMCs were available at data cut-off and IFNγ ELISpot and flow cytometry (in parentheses). N/A, not applicable. *8 and **7 for CD4^+^ T cell responses.

| Cohort | BNT162b2 vaccinated |  | Safety analysis |  | Antibody analysis |  |  |  |  |  |  | T-cell analysis |  |
| --- | --- | --- | --- | --- | --- | --- | --- | --- | --- | --- | --- | --- | --- |
|  | Prime | Boost | Day 1+ | Day 22±2+ | Day 1 | Day 8±1 | Day 22±2 | Day 29±3 | Day 43±4 | Day 50±4 | Day 85±7 | Day 1 | Day 29±3 |
| 1 µg | 12 | 11 | 12 | 11 | 12 | 12 | 12 | 11 | 10 | 10 | 0 | 9* (8) | 9 (8) |
| 10 µg | 12 | 11 | 12 | 11 | 12 | 12 | 11 | 11 | 11 | 11 | 11 | 9** (10) | 9 (10) |
| 20 µg | 12 | 12 | 12 | 12 | 12 | 12 | 12 | 12 | 12 | 12 | 10 | 9 (9) | 9 (9) |
| 30 µg | 12 | 12 | 12 | 12 | 12 | 12 | 12 | 12 | 11 | 12 | 12 | 10 (10) | 10 (10) |

**Extended Data Table 3a.** Summary of solicited local reactions.

| Time interval |  | 1 µg<br>(N=12) | 10 µg<br>(N=12) | 20 µg<br>(N=12) | 30 µg<br>(N=12) | Total<br>(N=48) |
| --- | --- | --- | --- | --- | --- | --- |
| Dose 1 up to day 7 after dose 1 | nn | 12 | 12 | 12 | 12 | 48 |
|  | Any local reaction, n (%) | 6 (50) | 12 (100) | 12 (100) | 10 (83) | 40 (83) |
|  | Any grade ≥3 local reaction, n (%) | 0 (0) | 0 (0) | 0 (0) | 0 (0) | 0 (0) |
| Dose 2 up to day 7 after dose 2 | nn | 11 | 11 | 12 | 12 | 46 |
|  | Any local reaction, n (%) | 4 (36) | 10 (91) | 10 (83) | 11 (92) | 35 (76) |
|  | Any grade ≥3 local reaction, n (%) | 0 (0) | 0 (0) | 0 (0) | 0 (0) | 0 (0) |
| Combined interval | nn | 12 | 12 | 12 | 12 | 48 |
|  | Any local reaction, n (%) | 7 (58) | 12 (100) | 12 (100) | 11 (92) | 42 (88) |
|  | Any grade ≥3 local reaction, n (%) | 0 (0) | 0 (0) | 0 (0) | 0 (0) | 0 (0) |

**Extended Data Table 3b.** Summary of solicited systemic reactions. The combined interval is the union of the intervals ‘Dose 1 up to day 7 after dose 1’ and ‘Dose 2 up to day 7 after dose 2’. N = number of participants in the analysis set; n = number of participants with the respective local (**a**) or systemic (**b**) reactions; nn = number of participants with any information on local (**a**) or systemic (**b**) reactions available.

| Time interval |  | 1 µg<br>(N=12) | 10 µg<br>(N=12) | 20 µg<br>(N=12) | 30 µg<br>(N=12) | Total<br>(N=48) |
| --- | --- | --- | --- | --- | --- | --- |
| Dose 1 up to day 7 after dose 1 | nn | 12 | 12 | 12 | 12 | 48 |
|  | Any systemic reaction, n (%) | 9 (75) | 12 (100) | 9 (75) | 9 (75) | 39 (81) |
|  | Any grade ≥3 systemic reaction, n (%) | 0 (0) | 0 (0) | 1 (8) | 0 (0) | 1 (2) |
| Dose 2 up to day 7 after dose 2 | nn | 11 | 11 | 12 | 12 | 46 |
|  | Any systemic reaction, n (%) | 4 (36) | 7 (64) | 10 (83) | 10 (83) | 31 (67) |
|  | Any grade ≥3 systemic reaction, n (%) | 0 (0) | 1 (9) | 1 (8) | 3 (25) | 5 (12) |
| Combined interval | nn | 12 | 12 | 12 | 12 | 48 |
|  | Any systemic reaction, n (%) | 9 (75) | 12 (100) | 11 (92) | 12 (100) | 44 (92) |
|  | Any grade ≥3 systemic reaction, n (%) | 0 (0) | 1 (8) | 2 (17) | 3 (25) | 6 (13) |

