## Supplementary Figures for "BNT162b2 induces SARS-CoV-2-neutralising antibodies and T cells in humans"

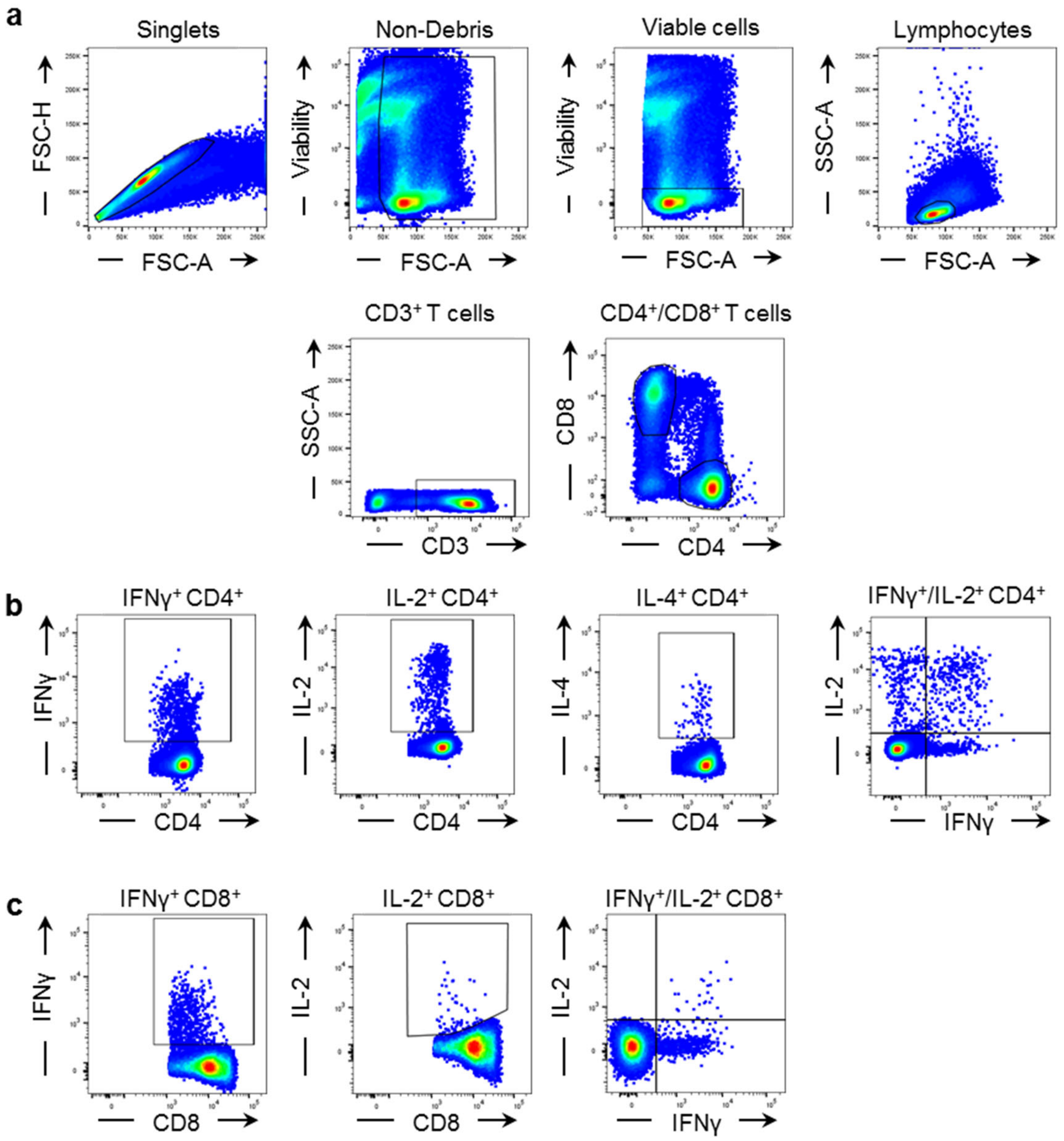


**Supplementary Figure 1 | Flow cytometry gating strategy for cytokine analysis by flow cytometry.**

Gating strategy for identification of IFNγ, IL-2 and IL-4 secreting T cells in PBMC samples. **a,** $CD4^+$ and $CD8^+$ T cells were gated within single, viable lymphocytes. **b, c,** Gating of IFNγ, IL-2 and IL-4 in $CD4^+$ T cells (**b**), and IFNγ and IL-2 in $CD8^+$ T cells (**c**).

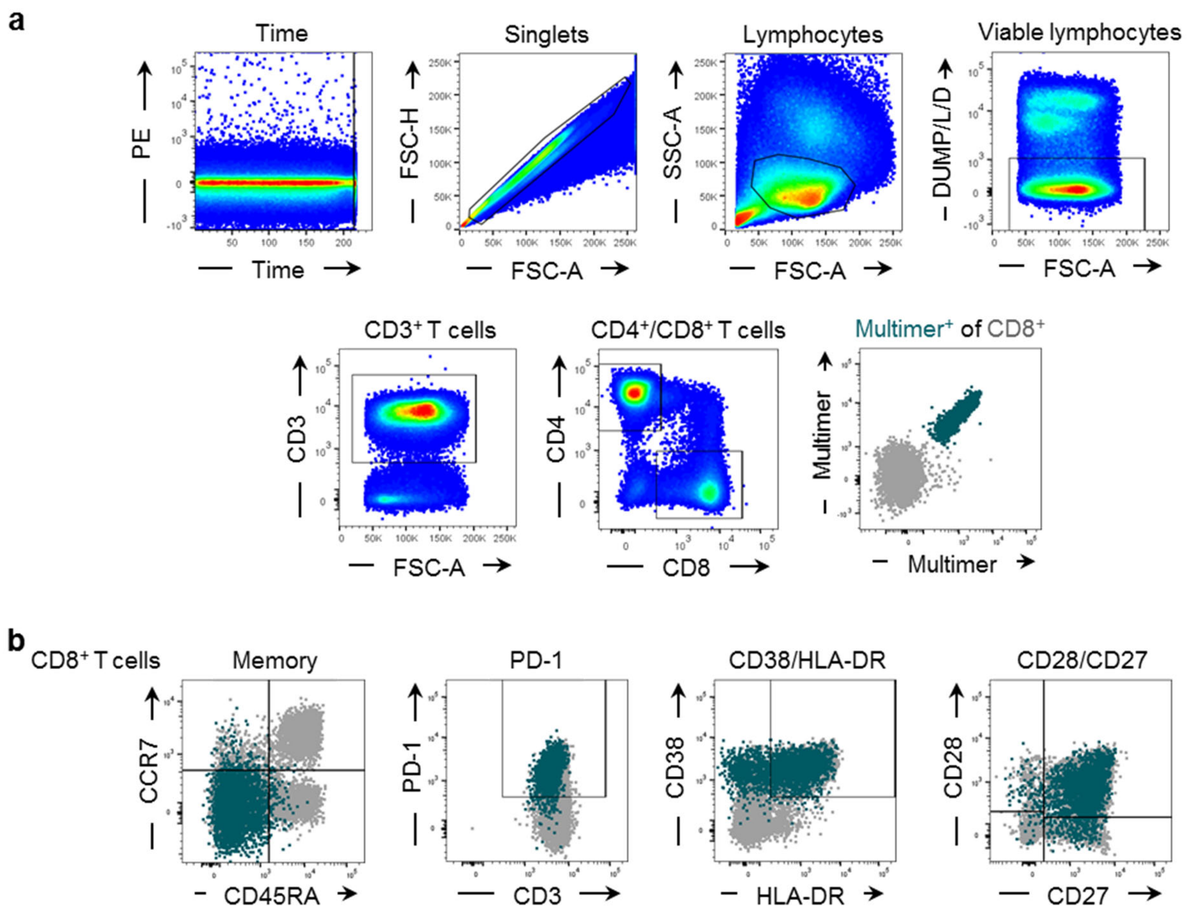


**Supplementary Figure 2 | Flow cytometry gating strategy for T cell specificity and subset analysis by flow cytometry.**

Gating strategy for identification and characterization of antigen-specific CD8$^+$ T cells in PBMC samples. **a,** Successive gates were applied to identify singlets, lymphocytes, DUMP (CD14, CD19, CD16 positive) and dead (L/D negative) cells, CD3$^+$ T cells, and CD4$^+$ or CD8$^+$ T cells. Antigen-specific CD8$^+$ T cells were gated as multimer double positive cells (green), with a combination of two fluorochromes labeling a defined MHC-epitope. **b,** Within CD8$^+$ T cells (grey), naïve (CD45RA$^+$/CCR7$^+$), central memory (CM; CD45RA$^-$/CCR7$^+$), effector memory (EM; CD45RA$^-$/CCR7$^-$) or effector (EMRA; CD45RA$^+$/CCR7$^-$) T cell subsets were gated, and activation status (CD38/HLA-DR), stages of differentiation (CD28/CD27) and expression of PD-1 determined. Multimer$^+$ CD8$^+$ T cells are highlighted in green.
